# IL-6R (trans-signaling) is a key regulator of reverse cholesterol transport in lipid-laden macrophages

**DOI:** 10.1101/2024.02.07.24302472

**Authors:** Fatema Al-Rashed, Halemah AlSaeed, Nourah Almansour, Fahd Al- Mulla, Yusuf A. Hannun, Rasheed Ahmad

## Abstract

**Background:** Atherosclerosis epitomizes a multifaceted cardiovascular disorder, predominantly characterized by the accumulation of cholesterol-laden plaques within arterial walls. Despite substantial research, the precise mechanisms governing the formation of these cholesterol-rich plaques remain partially elucidated. This study delves into the complex interplay of interleukin-6 (IL-6) receptors, shedding light on their pivotal role in orchestrating cholesterol homeostasis in human macrophages.

**Methods:** This investigation evaluated the correlation between interleukin-6 (IL-6), its receptors (IL6R/CD126), and glycoprotein 130 (gp130), alongside established atherosclerosis biomarkers. The cohort comprised 142 subjects, balanced between lean and obese individuals (71 each). Subsequent analyses utilized THP-1-derived macrophages to discern the biochemical repercussions of inhibiting IL-6 receptors on cellular mechanisms.

**Results:** Data indicates a significant upsurge in IL-6 secretion correlating with atherosclerotic manifestations in the obese subset, accompanied by a concomitant diminution in IL-6 receptors IL6R/CD126 and gp130 on circulating monocytes within this group. Pharmacological obstruction of the gp130 receptor in macrophages provoked pronounced alterations in lipid metabolism, notably impacting cholesterol management. These alterations were evidenced by an escalated expression of the LDLR gene, responsible for cholesterol uptake, and a surge in de novo cholesterol synthesis, marked by the upregulation of SREBF2 and its downstream effector, mevalonate kinase (MVK). Concurrently, an increase in HMG-CoA reductase protein levels was observed. Intriguingly, a rise in intracellular cholesterol production coupled with a reduction in ABCA1 levels was noted, suggesting a potential impediment in cholesterol efflux in cells deficient in gp130. This hypothesis was further substantiated by Filipin III staining, which indicated cholesterol retention in cells subjected to gp130 inhibition. Clinical implications of these discoveries were corroborated through experiments on PBMCs from lean participants, where the gp130 inhibitor curtailed cholesterol efflux to levels comparable to those in untreated cells.

**Conclusion:** Collectively, our research underscores the instrumental role of gp130 in the reverse cholesterol transport pathway of macrophages. These insights pave the way for novel therapeutic strategies targeting atherosclerosis and its associated cardiovascular complications, spotlighting gp130 as a potential focal point for intervention.

## Introduction

Atherosclerosis is a major cause of morbidity and death in developed countries(1). While chronic systemic inflammation is widely acknowledged across all stages of atherosclerosis, it is universally understood that this disorder primarily stems from lipid irregularities, with evidence from the buildup of oxidized low-density lipoproteins (oxLDL) in macrophages within the arterial walls (2). Current insights suggest that during the development of the atherosclerotic lesions, plasma circulatory monocytes migrate into the space beneath the endothelium. Here, they evolve into macrophages, ingesting altered lipoproteins, and eventually become foam cells (3). This process is believed to be instigated via cell scavenger receptors like SR-A and CD36 (4) or through the incorporation of cholesterol-bearing lipoprotein particles via the low-density lipoprotein receptor (LDLR) (5), which promotes the accumulation of lipids within their cellular structures and leads to the formation of foam cells. Another route to cholesterol-laden macrophages arises from the engulfment of apoptotic cells through efferocytosis (6). All in all, a substantial influx of cholesterol could potentially be harmful if the recycling of cholesterol within macrophages is disrupted (7, 8). It is interesting to point out that atherosclerosis is typically seen alongside other metabolic syndromes such as diabetes mellitus (DM), dyslipidemia and hypertension. Such observation strongly supports the vital impact of systemic inflammatory stimuli. Thus, a deeper understanding of the interaction between ox-LDL and inflammatory mediators involved in foam cell formation is critical to effectively prevent plaque rupture and the subsequent clinical life-threatening complications of atherosclerosis.

Interleukin-6 (IL-6) stands out as a notable cytokine within atherosclerotic tissue, with a substantial production originating from macrophages loaded with free cholesterol in advanced lesions (9). IL-6 receptors, including the primary IL-6 receptor (IL-6R) and the shared signal transducing subunit glycoprotein 130 (gp130), form a crucial duo in orchestrating cellular responses to interleukin-6 (IL-6)(10). The IL-6/gp130 complex activates the JAK/STAT3 pathway, triggering a cascade of events that regulate immune and inflammatory responses. It is noteworthy that IL-6 has been established as an independent risk factor for early coronary artery disease (11). Moreover, elevated levels of IL-6 are linked to an increased likelihood of myocardial infarction in healthy men (12). However, the precise role of IL-6 remains uncertain especially under different feeding habits, whether it acts as a causative factor, a consequence, or simply a marker of atherosclerosis. Indeed, experiments conducted on genetically modified mouse models predisposed to atherosclerosis reveal contradictory functions of IL-6 (13, 14). For instance, treating mice with recombinant mouse IL-6 resulted in larger lesions in both C57BL/6 and apoE-deficient mice fed a high fat/cholate diet (15). On the other hand, IL6-deficient mice developed larger fatty streak lesions compared to control mice when subjected to an atherogenic diet for 15 weeks (13).

To this end, the interplay between IL-6 receptors and secretion, especially in the cholesterol-rich inflammatory environment emblematic of atherosclerosis, is still under exploration. This study aims to discern the expression and correlation of IL-6 receptors (IL-6R/gp130) in human PBMCs isolated from both lean and obese subjects. Our findings indicate that in an obesity context, a spike in IL-6 secretion in media corresponds to an augmented cardiovascular risk, evident from the upregulation of total cholesterol. Conversely, the surface expressions of IL-6 receptors (IL6R/CD126) and gp130 were significantly diminished in obese participants. Delving deeper mechanistically using THP-1 transformed macrophages, we observed that the absence of the IL-6 receptor gp130 influenced macrophage cholesterol transport dynamics, amplifying cholesterol influx and curtailing cholesterol efflux, thus influencing macrophage cholesterol equilibrium favoring macrophage foaming.

## Materials and methods

### Study Design and Participant Recruitment

This investigation enlisted 142 individuals from a previously reported cohort (16, 17), all of whom were robust adults devoid of medical conditions, ensuring optimal physical health without any disabilities impeding movement. The study protocol entailed two distinct visits per participant. The initial encounter was dedicated to briefing the participants about the study specifics, potential risks, and subsequent procedures. Post briefing, participants who consented to partake in the study signed an informed consent form and underwent a comprehensive health screening.

### Data Collection and Assessment Procedures

Subsequent to the initial visit, participants were equipped with wearable accelerometers to monitor physical activity over a span of seven days. The follow-up visit encompassed comprehensive anthropometric and biomedical evaluations. The study adhered to a standardized protocol for all anthropometric measurements, employing consistent equipment to ensure uniformity. Height was accurately gauged using a portable stadiometer, with a precision of up to 1.0 cm, while weight measurements were ascertained to the nearest 0.2 kg using a standard physician’s beam scale (Detecto). The Body Mass Index (BMI) was computed using the standard formula: weight in kilograms divided by the square of height in meters (kg/m^2^).

### Biomedical Evaluations and Ethical Considerations

Post anthropometric assessment, participants proceeded to the phlebotomy unit for blood sample collection. Detailed physical and biochemical profiles of the participants, categorized by BMI, are delineated in Supplementary Table 1. The study was meticulously conducted in strict adherence to the ethical guidelines delineated in the Declaration of Helsinki. Ethical approval for the study was secured from the Kuwait Ministry of Health Ethical Board (Approval ID#: 1806/2021) and the Ethical Review Committee (ERC) of the Dasman Diabetes Institute, Kuwait (Approval ID#: RA MoH-2022-002), ensuring all participants provided written informed consent prior to their inclusion in the study.

### Isolation of human peripheral blood mononuclear cells (PBMC)

Peripheral blood mononuclear cells (PBMCs) were meticulously extracted from EDTA-treated vacutainer tubes collected from all study participants. The cells were isolated employing the Histo-Paque density gradient method in conjunction with Sepmate Tubes (StemCell Technologies, Vancouver, CN, Canada)(18). For the derivation of monocytic cells, the isolated PBMCs were plated in 6-well or 24-well plates (Costar, Corning Incorporated, Corning, NY, USA) at a density of 3×10^6 cells per well.

### Monocytic Cell Cultivation Conditions

The plated cells were cultured in a starvation medium consisting of RPMI-1640 culture medium (Gibco, Life Technologies, Grand Island, NY, USA), enriched with 2 mM glutamine (Gibco), 1 mM sodium pyruvate, 10 mM HEPES, 100 µg/mL Normocin, and 50 U/ml penicillin combined with 50 µg/mL streptomycin (P/S; Gibco). The incubation of the cells was conducted at a controlled temperature of 37°C, in a humidified atmosphere containing 5% CO2. After a period of 3 hours, non-adherent cells were gently removed to ensure the purity of the adherent monocytes. Subsequently, the adherent monocytes were rinsed with serum-free culture medium. The final cultivation phase involved a 24-hour incubation in RPMI supplemented with 2% fetal bovine serum, facilitating optimal cell growth and differentiation under these meticulously controlled conditions.

### Measurement of blood metabolic markers

The protocol for assessing blood metabolic markers was rigorously followed as delineated in prior research (19). Prior to the collection of blood samples for plasma isolation, essential physiological parameters, including blood pressure and heart rate, were accurately measured for each participant.

### Plasma Isolation and Biochemical Analysis

Post-collection, isolated plasma served as the primary medium for the quantitative analysis of various metabolic indicators. This included the measurement of triglyceride levels, total cholesterol, high-density lipoprotein (HDL)-cholesterol, and fasting glucose concentrations. To ensure the reliability and accuracy of the metabolic marker assessments, quality control sera were systematically employed throughout the testing procedures. This meticulous approach underscored the commitment to precision and the integrity of the data obtained from these critical biochemical measurements.

### Enzyme-Linked Immunosorbent Assay (ELISA) for Biomarker Detection

For the precise quantification of matrix metalloproteinase 9 (MMP9) and vascular endothelial growth factor (VEGF), the study employed commercially procured enzyme-linked immunosorbent assay (ELISA) kits (R&D Systems, Minneapolis, MN, USA). The assay procedures were meticulously executed in strict accordance with the manufacturer-provided protocols and guidelines established by previously published methodologies. This stringent adherence ensured the reliability and accuracy of the biomarker detection, fostering robust and replicable results within the study’s framework.

### Cell Culture and differentiation

#### Cultivation and Differentiation of Human Monocytic THP-1 Cells

Human monocytic THP-1 cells, acquired from the American Type Culture Collection (ATCC), were cultured in RPMI-1640 culture medium (Gibco, Life Technologies, Grand Island, USA). The culture medium was fortified with 10% fetal bovine serum, 2 mM glutamine, 1 mM sodium pyruvate, 10 mM HEPES, 100 µg/ml Normocin, and 50 U/ml penicillin, along with 50 µg/ml streptomycin (P/S; Gibco, Invitrogen, Grand Island, NY, USA). The cells were maintained under precise conditions at 37°C, in a humidified atmosphere supplemented with 5% CO_2_. For experimental purposes, the monocytic cells were seeded into 12-well plates (Costar, Corning Incorporated, Corning, NY, USA) at a density of 1×10^6^ cells per well, unless specified otherwise.

#### Macrophage Differentiation and Stimulation Protocol

Prior to stimulation, the monocytic cells underwent differentiation into macrophages using 10 ng/ml phorbol-12-myristate-13-acetate (PMA) over a period of three days. This was followed by a resting phase of an additional three days in serum-PMA-free RPMI media, ensuring the cells were fully primed for subsequent treatments. In the subsequent stimulation studies, differentiated macrophages were initially pre-treated with 10 ng/ml of SC144 (to inhibit the gp130 receptor), recombinant human IL-6 (hrIL-6), or a vehicle control for one hour. Post pre-treatment, the cells were stimulated with Human High Oxidized Low-Density Lipoprotein (oxLDL) (KB KALAN Biomedical; Cat no: 770252-4) for a duration of 24 hours, setting the stage for the exploration of various cellular responses and interactions under these defined conditions.

#### Real-Time Quantitative RT-PCR Process

The extraction of total RNA was conducted using the RNeasy Mini Kit (Qiagen, Valencia, CA, USA), strictly adhering to the manufacturer’s specified protocol. Following extraction, cDNA synthesis was performed utilizing 1 µg of total RNA with the aid of a high-capacity cDNA reverse transcription kit (Applied Biosystems, Foster City, CA, USA). The quantification of gene expression was carried out using the 7500 Fast Real-Time PCR System (Applied Biosystems, Foster City, CA, USA), coupled with the TaqMan® Gene Expression Master Mix (Applied Biosystems, Foster City).

#### Amplification and Data Normalization

For each reaction, 50 ng of cDNA was amplified using Inventoried TaqMan Gene Expression Assay products. The threshold cycle (Ct) values obtained were normalized against the housekeeping gene GAPDH to ensure accuracy in the relative quantification of the target mRNA. The relative quantities of mRNA, in comparison to control samples, were computed employing the ΔΔCt-method, with the expression levels presented as fold-change relative to the mean expression of the control gene. The baseline expression level for the control treatment was set at 1. Data are represented as mean ± SEM. The threshold for statistical significance was established at p < 0.05. For a comprehensive overview of the primers utilized in the study, refer to Supplementary Table 2, where the primer list is extensively detailed.

#### Western Blotting Procedure

For Western blot analysis, cells were first collected and then incubated with lysis buffer (comprising Tris 62.5 mM at pH 7.5, 1% Triton X-100, and 10% glycerol) for a duration of 30 minutes. Post-incubation, the cell lysates underwent centrifugation at 14,000x g for 10 minutes, facilitating the separation and subsequent collection of the supernatants. The protein concentration within these lysates was quantitatively assessed using the Quickstart Bradford Dye Reagent, 1x Protein Assay kit (Bio-Rad Laboratories, Inc, CA).

#### Sample Preparation and Gel Electrophoresis

Protein samples (50 µg in 20 µl) were prepared by mixing with sample loading buffer and then heated for 5 minutes at 95°C. A quantity of 12.5 µg from these prepared samples was loaded and subjected to electrophoretic separation on 12% polyacrylamide gels using SDS-PAGE. Subsequently, the separated cellular proteins were transferred onto Immuno-Blot PVDF membranes (Bio-Rad Laboratories, USA) through electroblotting.

#### Membrane Processing and Visualization

The membranes were initially blocked using 5% non-fat milk in PBS for one hour to prevent non-specific binding. This was followed by incubation with various primary antibodies (detailed in Supplementary Table 3). After primary antibody incubation, the membranes were washed four times with TBS and then incubated for 2 hours with species-specific HRP-conjugated secondary antibodies (Promega, Madison, WI, USA). The immunoreactive bands were visualized through the use of the Amersham ECL plus Western Blotting Detection System (GE Health Care, Buckinghamshire, UK) and captured using the Molecular Imager ® ChemiDoc^TM^ MP Imaging Systems (Bio-Rad Laboratories, Hercules, CA, USA), ensuring clear and precise documentation of the protein expression patterns.

#### Cholesterol Efflux Assay Implementation

The measurement of cholesterol efflux was rigorously performed in accordance with the protocol specified by the Cholesterol Efflux Assay Kit (Cell-based) (abcam; ab196985). This involved the incubation of either differentiated cells or isolated PBMCs with labeling and equilibrium mediums. Following this preparatory step, the cells underwent a treatment phase with cholesterol acceptors for a period of four hours, utilizing LDL/VLDL-depleted human serum as the specific cholesterol acceptor for the assay.

#### Fluorescence Quantification and Efflux Percentage Calculation

Post-treatment, the fluorescence emanating from the cell lysate was measured at the wavelengths of 485/523 nm. This fluorescence measurement is critical as it reflects the extent of cholesterol that has been effluxed from the cells. To accurately quantify the percentage of cholesterol efflux, the following formula was applied:

### % Cholesterol Efflux = (RFU of Supernatant)/ (RFU of Cell Lysate + RFU of Supernatant) x 100

#### Flow Cytometry—Staining of Cell-Surface Markers

To determine the expression of IL-6 receptors; IL-6Ra/ CD126 and gp130/CD130 in circulatory monocytes, flow cytometry analysis was conducted using the isolated PBMCs. The cells were first resuspended in FACS staining buffer (BD Biosciences) and subjected to a blocking phase with human IgG (Sigma; 20 µg) for 30 minutes on ice. Post-blocking, the cells underwent a washing step, followed by resuspension in 100 µL of FACS buffer.

#### Staining and Incubation Procedures

For the staining process, the cells were treated with mouse anti-human CD14-PE-Cy™7 (cat# 560919, BD Pharmingen™), mouse anti-human CD126-APC (cat# 562090, BD Pharmingen™), and mouse anti-human CD130-PE (cat# 555757, BD Pharmingen™). The stained cells were incubated on ice for a total of 30 minutes, with gentle mixing at 10-minute intervals to ensure uniform staining. This was followed by three washes with FACS buffer and a final fixation step, involving resuspension in 2% paraformaldehyde solution.

#### Intracellular Staining and FACS Analysis

For intracellular staining, cells were treated with fixation/permeabilization buffer (Cat# 00-5523-00, eBioscience, San Diego, CA, USA) for 20 minutes at 4°C, then washed and stained with intracellular inflammatory cytokine rat anti-human IL-6-PE (cat# 554545, BD Pharmingen™). Following staining, the cells were washed again and resuspended in PBS supplemented with 2% FCS for subsequent FACS analysis (FACSCanto II; BD Bioscience, San Jose, USA). Data analysis was executed using BD FACSDiva™ Software 8 (BD Biosciences, San Jose, USA). To accurately set the thresholds for negative versus positive gates, non-stain cells and isotype-specific respective control antibodies were employed.

#### Oil-Red-O Lipid Staining Procedure

The visualization of lipid accumulation in treated cells was achieved through the application of Oil-Red-O lipid staining, a technique renowned for its specificity to lipids, primarily triglycerides and lipoproteins.

#### Preparation and Fixation of Cells

Initially, the treated cells were gently washed with PBS to remove any residual media or non-adherent cells. This was followed by a fixation process, where the cells were immersed in 10% formalin solution for a duration of 30 minutes, ensuring the preservation of cellular structures and lipid content.

#### Staining and Visualization Process

Post fixation, the cells were subjected to a series of washes with ddH2O to remove the fixative. Subsequently, the cells were prepared for staining by incubating them in 60% isopropanol for 5 minutes. This step enhances the permeability of the cell membrane, facilitating the effective penetration of the Oil Red O stain. The Oil Red O working solution, freshly prepared and reconstituted with water, was then added to the cells and allowed to incubate for 20 minutes. During this period, the lipid components within the cells selectively absorbed the stain. Following the incubation, the excess stain was carefully discarded, and the cells were thoroughly washed with ddH2O to remove any non-specifically bound stain. and wre considered ready for visualization under the microscope.

### Filipin cholesterol staining

Treated cells were fixed using 4% paraformaldehyde, rinsed, and then incubated with Filipin complex (50 μg/ml) for two hours at room temperature, protected from light. Post-staining, samples were washed and mounted for fluorescence microscopy. Imaging was performed using a UV filter set to specifically capture Filipin’s fluorescence with excitation (355 nm) and emission (420 nm) through a standard UV filter block. Image acquisition was standardized to minimize photo-oxidation and fading.

### BODIPY neutral lipid staining

For the detection of neutral lipids, BODIPY 493/503 was used. After fixing with 4% paraformaldehyde, cells were stained with BODIPY 493/503 at a concentration of 2μM for 15 minutes at room temperature in the dark. Following staining, cells were washed and mounted for imaging. We used fluorescence microscopy with the appropriate filters to detect BODIPY’s green fluorescence at excitation of (493nm) and emission of (503 nm), carefully calibrating the settings to prevent photobleaching.

### Phalloidin f-actin staining

Phalloidin stain (6.6 µM) diluted with PBS used to incubate the cells for 20min at room temperature, then the cells are visualized under fluorescent microscopy.

#### Confocal Microscopy Imaging Protocol

##### Instrumentation and Configuration

Imaging was conducted using an inverted Zeiss LSM710 spectral confocal microscope (Carl Zeiss, Gottingen, Germany), paired with an EC Plan-Neofluar 40×/1.30 oil DIC M27 objective lens. The samples underwent excitation utilizing a dual-laser system comprising a 543 nm HeNe laser and the 405 nm line of an argon ion laser, ensuring optimal illumination for each specific fluorophore. Following excitation, the emission detection bandwidths were optimized and configured using the Zeiss Zen 2010 control software. This careful calibration was crucial for accurately capturing the fluorescence emissions, thereby allowing for precise and detailed visualization of the cellular structures and components under study.

##### Statistical Methodology and Data Presentation

The statistical analyses were executed using GraphPad Prism software (La Jolla, CA, USA). Data are presented as mean ± standard error of the mean (SEM) unless specified otherwise.

##### Normality Testing and Group Comparison Methods

Initial data examination included the Shapiro–Wilk test to confirm normal distribution. For comparative analyses, group means were contrasted using two-tailed tests for parametric data and Wilcoxon–Mann– Whitney U tests for non-parametric datasets, ensuring appropriate and robust statistical inference. Additionally, for assessments involving three groups, one-way ANOVA and exact Kruskal–Wallis tests were employed for parametric and non-parametric data respectively, catering to the dataset’s distribution characteristics.

##### Correlation Analysis and Significance Thresholds

Correlation strength and direction between two variables were quantified using Pearson’s correlation coefficient “r” for parametric data, and Spearman’s correlation coefficient for non-parametric datasets. The significance threshold was set at P < 0.05. The study further delineates levels of significance with notations: ns (non-significant), *P < 0.05, **P<0.01, ***P< 0.001, and ****P < 0.0001, in comparison to control, facilitating a nuanced understanding of the statistical findings in relation to the control group.

## Results

### Obese individuals exhibit higher IL-6 plasma levels and reduced expression of IL-6 receptors on circulatory monocytes

To begin investigating the potential role of IL-6 receptors in modulating human macrophages’ maintenance of cholesterol homeostasis, we first assessed the secretion of IL-6 cytokine in our cohort. Similar to previous findings, the plasma secretion of IL-6 in our cohort was found to be significantly elevated in obese individuals compared to lean participants (**Figure 1A**). This observed elevation in IL-6 secretion was also observed to be parallel with the increase in atherosclerosis indicators, including total cholesterol (**Figure 1B**), triglycerides (TG) (**Figure 1C**), and C-reactive protein (CRP) (**Figure 1D**), while also aligning with the reduction in high-density lipoprotein (HDL) levels (**Figure 1E**). Further correlation analysis showed a significant positive association ofIL-6 plasma secretion with adverse atherosclerotic markers including total cholesterol, TG and CRP, whereas as expected a significant negative association with HDL (**Figure 1F-1I**).

**Figure 1.**
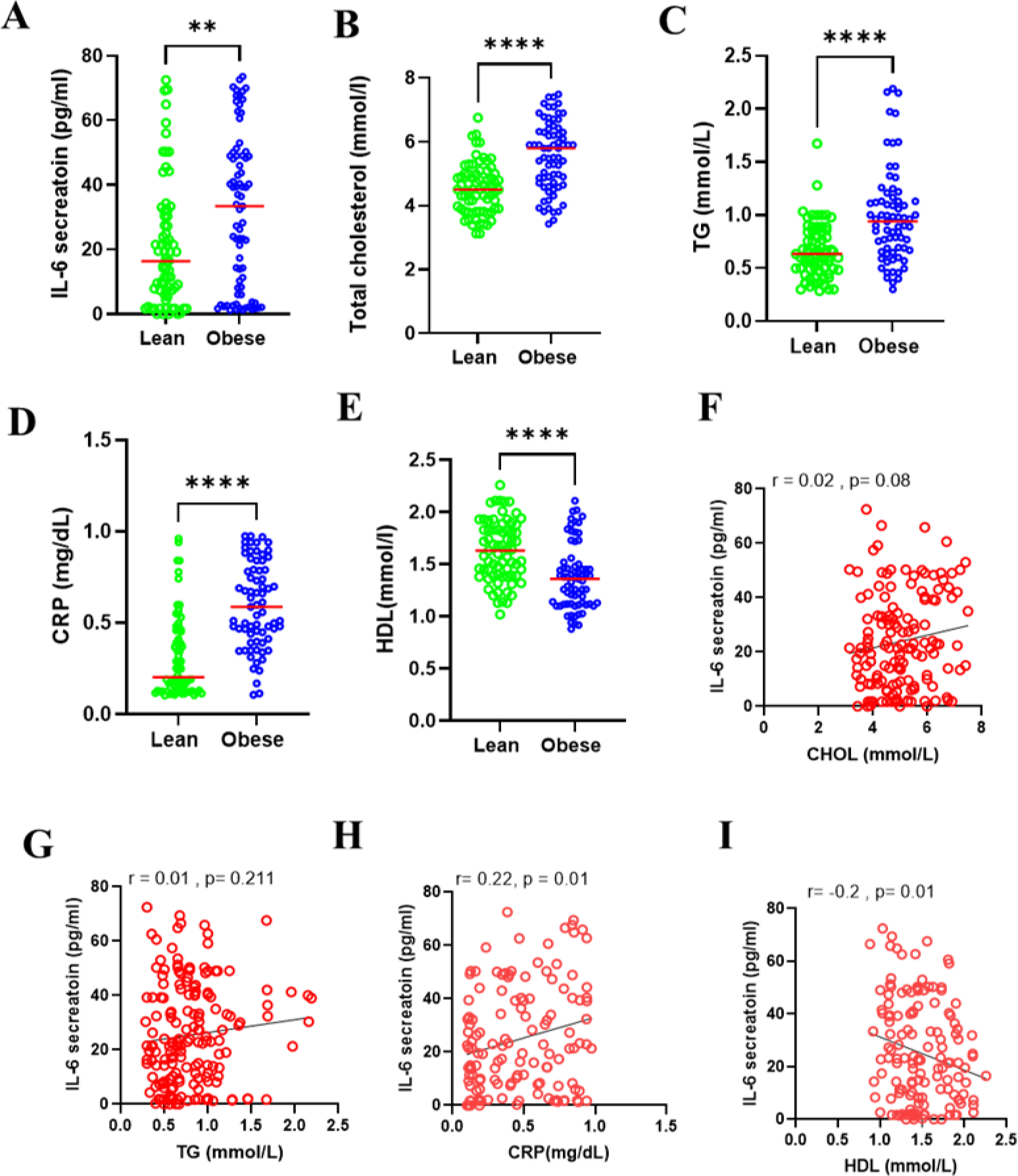
IL-6 secretion is elevated in obese individuals’ plasma and is associated with atherosclerosis related markers. Plasma levels of atherosclerosis related cytokines secretion of (A) IL-6, (B) Total cholesterol, (C) TG, (D) CRP, and (E) HDL. Two-tailed /Wilcoxon–Mann–Whitney U tests were used to assess the differences between two Lean vs Obese. Pearson’s/Spearman’s correlation coefficient and linear regression analysis was conducted to investigate the relationship between secreted IL-6 cytokines and atherosclerosis related markers (F-I). All data are expressed as mean ± SD. P ≤ 0.05 was considered statistically significant (**P < 0.01, ****P < 0.0001).

However, further investigation on the expression and activity of IL6 receptor IL6R/CD126 through flow cytometry defined a different dynamic, as it was observed that lean participants presented a significantly higher representation of CD14^+^CD126^+^ expression on their circulatory monocyte surface (**Figure 2A**). This expression was positively associated with elevated HDL levels in the plasma (**Figure 2B**) and negatively correlated with lower systolic and diastolic blood pressure in participants (**Figure 2F and 2G**). However, no significant association was found to other measured atherosclerotic markers including TG, total cholesterol and heart rate (HR) (**Figure 2C, 2D, and 2E**). We also did not observe any association between IL-6R surface expression and atherosclerotic inflammatory markers CRP, Matrix metallopeptidase 9 (MMP9) or Vascular endothelial growth factor (VEGF) (**Figure 2H-2J**).

**Figure 2:**
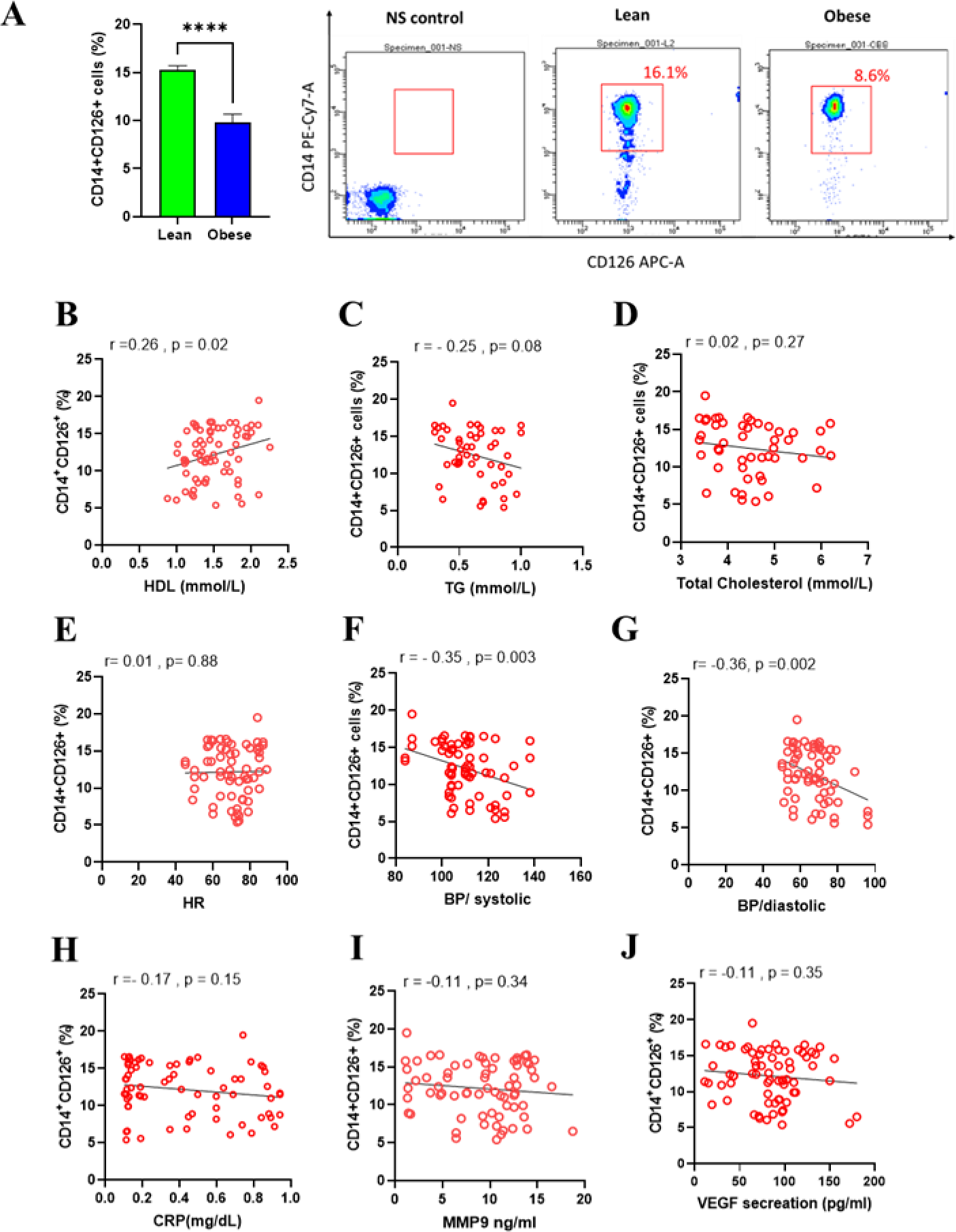
IL-6 receptors expression is markedly decreased in obese individuals’ circulatory monocytes. PBMCs were isolated from human blood samples obtained from lean and obese individuals. (A) Bar-graph of Flow cytometry analysis of CD14+CD126+ subsets percentage, (B) Representative dot-plot analysis of CD14+CD126+ subsets expression (B-J) Correlation between CD14+CD126+ subset percentages and atherosclerosis related markers. Data are presented as mean ± SD. *p≤0.05, **p≤0.01, ***p≤0.001, ****p≤0.0001.

Interestingly, when we measured the expression levels of the membrane glycoprotein 130 (gp130) transducing chain through flowcytometry analysis of CD130 surface marker, we encountered a higher abundance in all participants with a similar pattern of elevated CD126 expression observed in lean participants compared to those with obesity (**Figure 3A**). Unlike CD126, the elevated expression levels of gp130 were found to be significantly associated with further favorable lipid markers for atherosclerosis. Elevated gp130 expression was positively correlated with higher HDL (**Figure 3B**) and negatively associated with total cholesterol and TG (**Figure 3C and 3D**). The elevation of gp130 expression was also found to be associated with significantly reduced systolic and diastolic blood pressure with no impact on HR (**Figure 3E-3G).** Most importantly, gp130 expression levels was significantly found to be associated with all measured atherosclerotic inflammatory markers (**Figure 3H-3J),** raising the possibility of a prominent role of gp130 in activating the downstream inflammatory signal for these observed markers.

**Figure 3:**
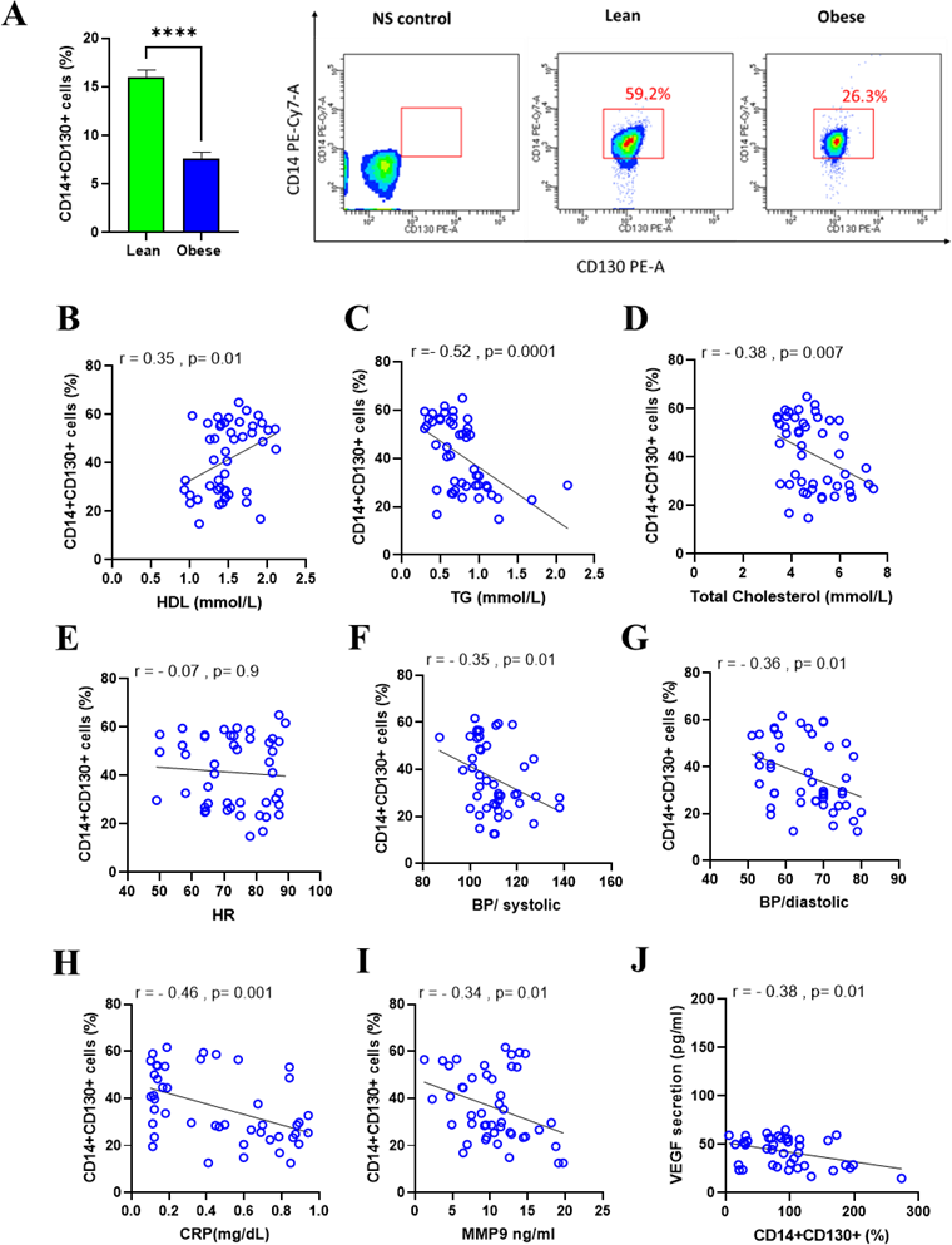
IL-6 trans-membrane receptor gp130 is markedly decreased in obese individuals’ circulatory monocytes. PBMCs were isolated from human blood samples obtained from lean and obese individuals. (A) Bar-graph of Flow cytometry analysis of CD14^+^CD130^+^ subsets percentage, (B) Representative dot-plot analysis of CD14^+^CD130^+^ subsets expression (B-J) Correlation between CD14^+^CD130^+^ subset percentages and atherosclerosis related markers. Data are presented as mean ± SD. *p≤0.05, **p≤0.01, ***p≤0.001, ****p≤0.0001.

Overall, these results indicate that elevated IL-6 secretion was associated with atherosclerotic indicators in obese individuals, while decrease in IL-6R and gp130 expression on circulatory monocytes was noted in the same cohort. Increased gp130 expression positively correlated with HDL and negatively correlated with total cholesterol, TG in lean individuals indicating a central function of IL-6R and gp130 in controlling pathology of atherosclerosis.

### Pharmaceutical inhibition of gp130 receptor induces lipid accumulation in macrophages

The role of the JAK/STAT3 pathway downstream of gp130 signaling is well documented to promote atherosclerosis development (20). To define the role of this observed decrease in gp130 in atherosclerosis at the molecular level, we treated macrophages derived fromTHP-1 cells with 10ng/ml of the gp130 inhibitor SC144 in the presence and absence of oxLDL. The inhibition of the gp130 receptor had a significant impact on the level of intracellular lipid accumulation within the macrophages; this impact was further enhanced in the presence of oxLDL, showing positive cytoplasmic granularity and neutral lipid accumulation measured through oil red O staining (**Figures 4A and 4B**). The influence of SC144 on lipid accumulation was further supported by the increase in PLIN2 gene expression (**Figure 4C**), which advocates the formation of lipid droplets under gp130 inhibition. Furthermore, we aimed to delve deeper into understanding the impact of gp130 inhibition on lipid accumulation by examining the expression of key genes recognized for their control over various facets of lipid metabolism. The inhibition of gp130 significantly upregulated low-density lipoprotein receptor (LDLR) gene expression regardless of oxLDL supplementation (**Figure 4D**). Interestingly, no statistically significant alteration was observed in CD36 gene expression (**Figure 4E**), reinforcing the specificity of the observed effects on cholesterol regulation.

**Figure 4:**
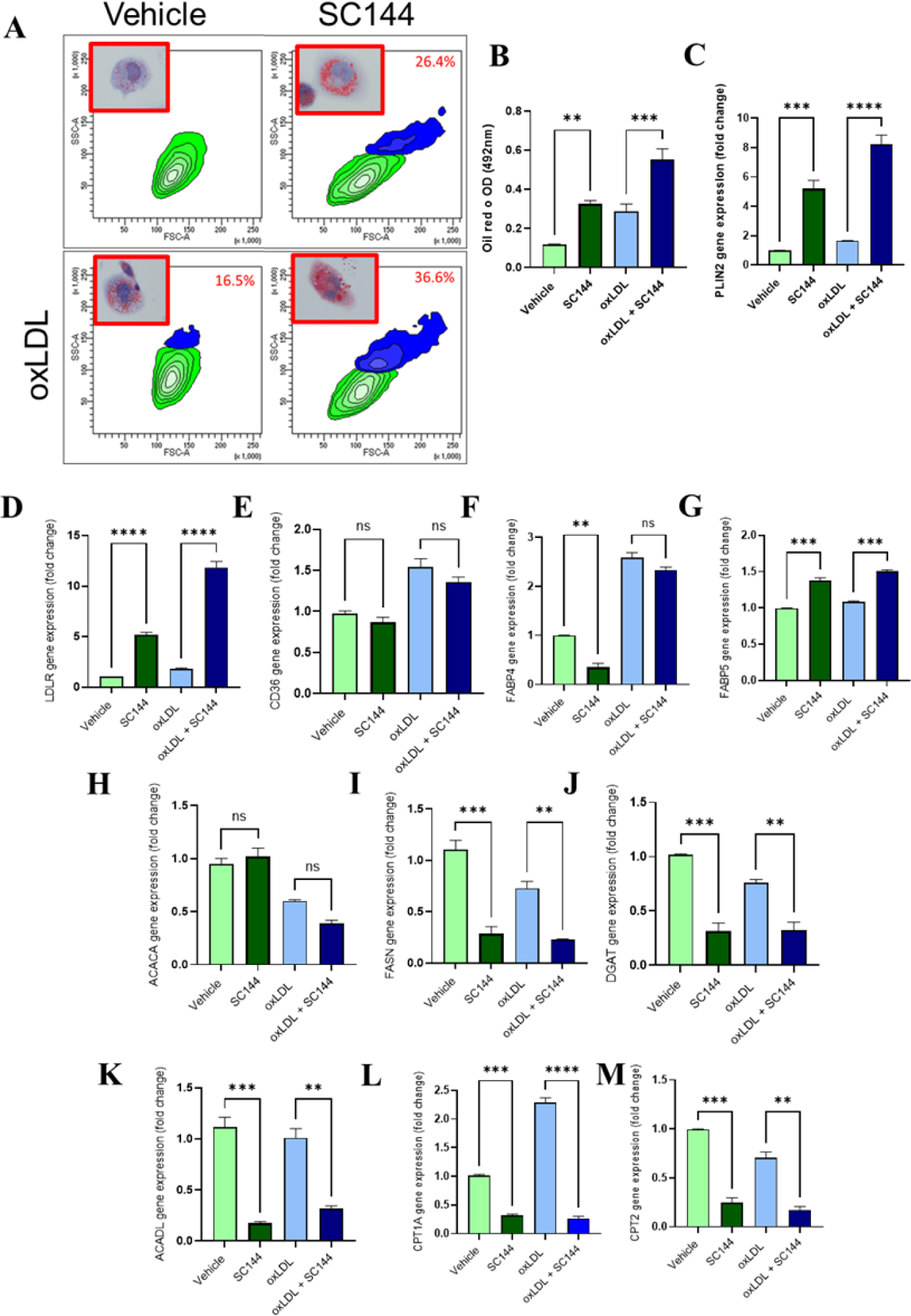
Pharmaceutical inhibition of gp130 receptor induces lipid accumulation in macrophage transformed THP-1 cells. THP-1-derived macrophages were pre-treated with gp130 inhibitor or vehicle control followed by stimulation with OxLDL (150uM; Thermo # L34357). (A) Representative dot plot of cellular granulation of macrophages with representative image of Oil red O staining. (B) Smi - quantitative analysis of Oil red O extraction. (C) Gene expression analysis of PLIN2. (D) Gene expression of LDLR. (E) Gene expression of CD36. (F) Gene expression of FABP4. (G) Gene expression of FABP5. (H) Gene expression of ACACA. (I) Gene expression of FASN. (J) Gene Expression of DGAT. (K) Gene Expression of ACADL. (L) Gene expression of CPT1A and (M) Gene expression of CPT2. Data are presented as mean ± SEM values (n=3-4) and compared between groups using one-way ANOVA with Tukey’s multiple comparisons test. *p≤0.05, **p≤0.01, ***p≤0.001, ****p≤0.0001.

Furthermore, our findings present a remarkable reduction in expression of Fatty Acid Binding Protein 4 (FABP4), underscoring the inhibitor’s capacity to modulate fatty acid shuttling across the cytoplasm. However, under the influence of oxLDL supplementation, this effect was nullified, giving rise to a pervasive elevation evident in both the vehicle and SC144 test groups (**Figure 4F**). Interestingly, an opposite observation was demonstrated with FABP5 as it showed a significant elevation in its expression under both oxLDL presence and absence (**Figure 4G**).

No Significant change in the gene expression of Acetyl-CoA Carboxylase Alpha (ACACA), which serves as the rate-limiting catalyst for long-chain fatty acid biosynthesis (**Figure 4H**). In contrast, similar to oxLDL treated group, a notable reduction was observed under SC144 treatment in comparison to their respective vehicle control regardless of oxLDL presence. This observation was found in the gene expression of Fatty Acid Synthase (FASN), a downstream component of ACACA (**Figure 4I**), as well as Diacylglycerol O-Acyltransferase (DGAT), a pivotal enzyme in the metabolic pathway essential for triacylglycerol production (**Figure 4J**). Furthermore, gp130 inhibition induced significant reduction in β-oxidation, since the expression of Acyl-CoA Dehydrogenase Long Chain (ACADL) and both Carnitine palmitoyl transferase 1/2 (CPT1A and CPT2) were reduced compared to controls regardless to oxLDL treatment as well (**Figure 4K to 4M**). Under gp130 inhibition, the observed profile includes macrophage foaming, elevated expression of cholesterol regulatory uptake receptors and fatty acid transportation, and reduced de novo lipogenesis, triglyceride production, and liberation. This suggests an association with increased lipid influx and utilization, reflecting the cell’s effort to regulate its response and prevent further uptake and synthesis of fatty acids in the absence of the gp130 receptor. These findings potentially contribute to shifts in lipid metabolism and related pathways, primarily within the realm of cholesterol metabolism.

### Pharmaceutical inhibition of gp130 receptor modulates cholesterol homeostasis program in macrophages

In order to gain deeper insights into the role of the gp130 receptor in macrophage cholesterol homeostasis, we conducted a comprehensive investigation into the impact of the gp130 inhibitor SC144, both in the presence and absence of oxLDL, on a select subset of key genes involved in cholesterol homeostasis. Similar to our previous analysis, we employed quantitative polymerase chain reaction (qPCR) to assess the expression levels of these genes, allowing us to elucidate the molecular mechanisms underlying cholesterol regulation in macrophages. Our initial findings revealed that the inhibition of the gp130 receptor did not elicit a significant alteration in the expression of the Sterol Regulatory Element-Binding Transcription Factor 1 (SREBF1) gene when compared to the vehicle control. Interestingly, however, both vehicle-treated and SC144-treated macrophages exhibited a reduction in SREBF1 expression upon oxLDL stimulation (**Figure 5A**). This observation suggests that oxLDL plays a pivotal role in modulating the expression of this gene, irrespective of gp130 inhibition. Remarkably, a noteworthy trend was observed in the expression of SREBPF2 under conditions of gp130 inhibition, although this increase did not reach statistical significance. Of particular interest was the finding that following oxLDL stimulation, the expression of SREBF2 remained significantly elevated in the presence of gp130 inhibition whereas oxLDL-treated vehicle cells exhibited a significant drop in expression compared to non-stimulated vehicle cells (**Figure 5B**). This suggests that gp130 inhibition may contribute to the sustained upregulation of SREBF2 in response to oxLDL.

**Figure 5:**
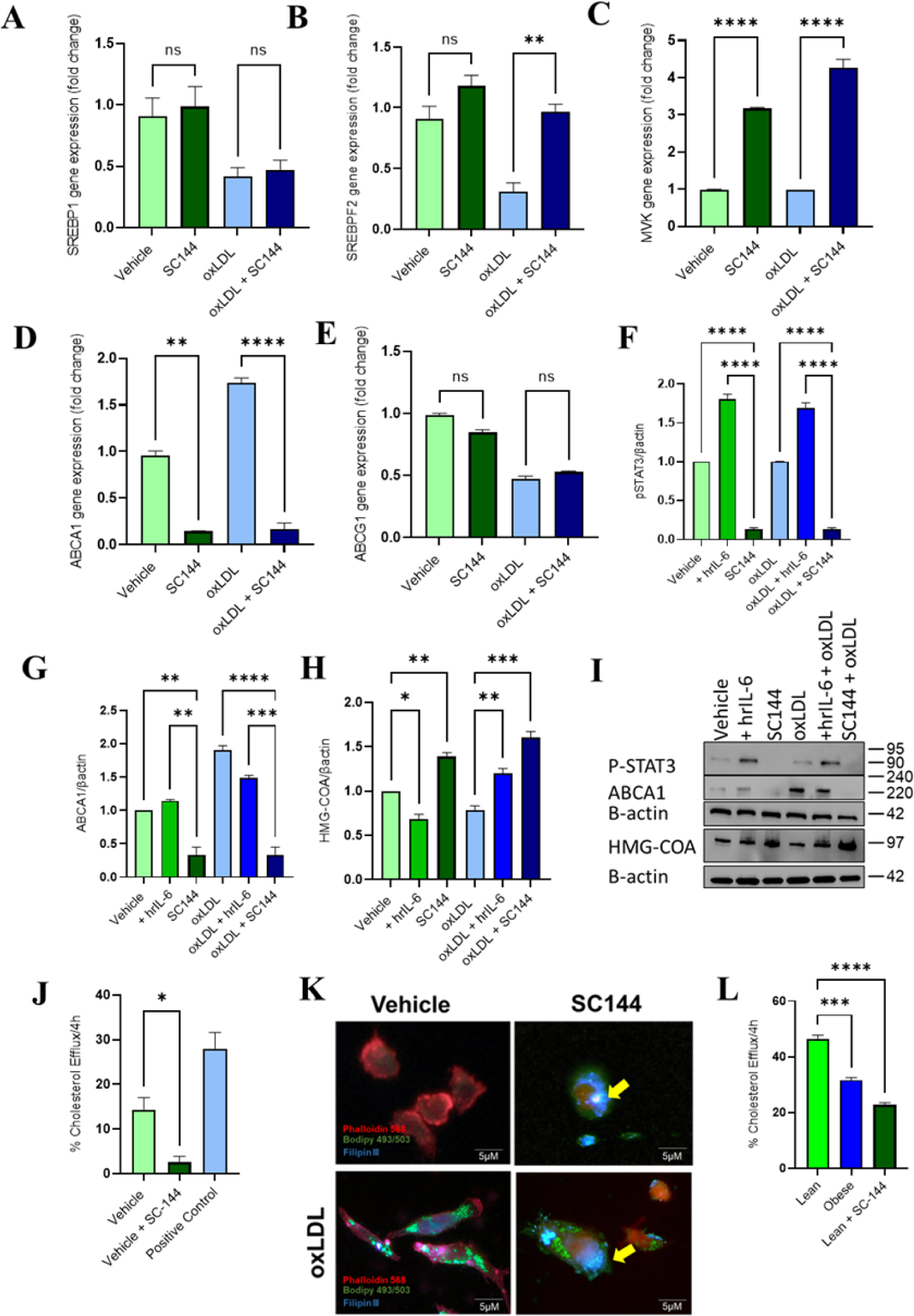
Pharmaceutical inhibition of gp130 receptor modulates cholesterol homeostasis in macrophage transformed THP-1 cells. THP-1-derived macrophages were pre-treated with gp130 inhibitor or vehicle control followed by stimulation with OxLDL (150uM; Thermo # L34357). QRT-PCR analysis was conducted for (A) SREBP1 gene expression. (B) SREBP2 gene expression. (C) MVK gene expression. (D) ABCA1 gene expression. (E) ABCG1 gene expression. Western Blot analysis for protein expression for (F) phospho-STAT3 corrected over β-actin. (G) ABCA1 corrected over β-actin. (H) HMG-COA corrected over β-actin. (I) Representative immune blots for protein expression. (J) Cholesterol efflux assay conducted for THP-1 transformed macrophages treated with SC144. (K) Immun-floresence microscopy images of treated THP-1 transformed macrophages stained with filipin III cholesterol stain (Blue), BODIPY 493/503 for neutral lipids (Green) and Phalloidin to labeled actin filaments (red) (Scale Bar 5µM). (L) Cholesterol efflux assay conducted on isolated and treated PBMC from both Lean and Obese individuals. Data are presented as mean ± SEM values (n=3-4) and compared between groups using one-way ANOVA with Tukey’s multiple comparisons test. *p≤0.05, **p≤0.01, ***p≤0.001, ****p≤0.0001.

The impact of SREBF2 gene expression was further reflected downstream in the expression of Mevalonate Kinase (MVK). Notably, a similar trend of significant upregulation in cholesterol biosynthesis was observed under conditions of gp130 inhibition, regardless of oxLDL stimulation (**Figure 5C**). These findings indicate a role for gp130 in regulating cholesterol biosynthesis, particularly through the modulation of SREBPF2 and its downstream targets. Additionally, we examined the expression of ATP Binding Cassette Subfamily A Member 1 (ABCA1) and G Member 1 (ABCG1), both of which play crucial roles in cholesterol efflux and are regulated by SREBP2. The inhibition of gp130 resulted in a remarkable suppression of ABCA1 gene expression, which was not rescued even under oxLDL stimulation (**Figure 5D**). Conversely, no significant difference was detected in ABCG1 gene expression under SC144 treatment compared to the vehicle control, with oxLDL stimulation leading to a reduction in gene expression regardless of gp130 inhibition (**Figure 5E**). These findings highlight the distinct regulatory roles of gp130 in controlling these efflux-related genes. To further verify this observation, we explored whether gp130 inhibition could affect the total protein levels of cholesterol efflux protein ABCA1 and the cholesterol *de novo* synthesis enzyme HMG-CoA. Treatment with SC144 significantly inhibited the activation of the gp130 receptor, thereby suppressing the downstream signaling cascade, including the phosphorylation of STAT3. Consistent with our mRNA observations, our protein analysis indicated a reduction in ABCA1 protein expression, while HMG-CoA expression was significantly increased, suggesting a potential shift toward cholesterol retention (**Figure 5F-5I**).

We then evaluated the functional significance of these changes through the use of a cholesterol efflux assay, and the results showed that SC144 treatment under oxLDL stimulation significantly increased cellular cholesterol levels by reducing its efflux to the media (**Figure 5J**). Intriguingly, we further examined the subcellular localization of cholesterol by staining with filipin to visualize cholesterol deposits (depicted in blue) and BODIPY 493/503 staining to visualize neutral lipids (depicted in green). Under conditions of gp130 inhibition, we observed an enhanced filipin signal, which partially colocalized with areas positive for neutral lipids (**Figure 5K**). While the semiquantitative nature of this staining necessitates further analyses to elucidate the dynamics of cholesterol and neutral lipids within the plasma membrane after gp130 inhibition, our observations suggest a notable accumulation of intracellular cholesterol in the treated group. In an attempted to translate this observation to a human *ex-vivo* model, we assessed the cholesterol efflux capacity of peripheral blood mononuclear cells (PBMCs) isolated from both lean and obese individuals. Furthermore, PBMCs obtained from lean subjects were pre-treated with SC-144, simulating the gp130 inhibitory milieu observed in obese participants. Fascinatingly, PBMCs from lean individuals exhibited superior cholesterol efflux capabilities compared to those from obese counterparts. Notably, upon gp130 inhibition, the cholesterol efflux potential of these cells was markedly attenuated, aligning closely with patterns observed in obese-derived PBMCs (**Figure 5L**). Collectively, our findings pertaining to gene and protein expression profiles underscore the involvement of cholesterol metabolism, particularly through pathways related to cholesterol synthesis and uptake, as influenced by gp130 inhibition. However, this increase in cholesterol synthesis is not matched by an equivalent increase in cholesterol clearance, as evidenced by the reduction in ABCA1 expression. These results shed light on the intricate interplay between gp130 signaling and macrophage cholesterol homeostasis.

## Discussion

Obesity is characterized as a chronic inflammatory condition, and it is considered a major risk factor for the development of atherosclerosis. Under obesity setting, the elevation of several macrophage-induced chemokines and cytokines has been promoted as a key mediator of local and systemic inflammation, oxidative stress, and abnormal lipid metabolism within the arterial walls. Under these contacts, the role of IL-6 cytokine specifically in the development of atherosclerosis has long been debated, as its elevation in atherosclerotic patient plasma has been reported by several groups. Regardless, the direct impact of circulatory IL-6 on the progression of atherosclerosis remains not well defined. In this work, we aimed to characterize IL-6 cytokine secretion and IL-6R/gp130 receptor signaling in both lean and obese population and correlated our findings with atherosclerosis risk markers. Our findings provide compelling insights into the molecular intricacies of atherosclerosis, particularly the role of IL-6 receptors in modulating human macrophage cholesterol homeostasis. The marked decrease in IL-6 receptors expression on circulatory monocytes in obese individuals, specifically gp130 despite elevated IL-6 secretion, suggests a complex regulatory mechanism that requires intensive further investigation. Similar to IL-6 secretion, the role of gp130 under inflammatory setting seemed to be diverse. In a study conducted by *Duan et al,* (21) it was demonstrated that CD146 interaction with gp130 and the subsequent activation of its signal is a major promoter of adipocyte infiltrated macrophages and a driven trigger for these macrophages to polarize toward a pro-inflammatory phenotype. However, this activation of pro-inflammatory response downstream of gp130 is also debated upon its protective outcome in part, by increasing the expression of suppressor of cytokine signalling-3 (SOCS3) (22, 23).Regardless, the role of gp130 expression seems to play a different role in metabolic diseases such as diabetes, obesity, and atherosclerosis. In a work presented by *Carbonaro et al*, it was shown that by restoring hepatic IL-6-gp130 signaling pathway through constitutive activation of gp130 substantially reduced lipid accumulation in human hepatocytes thus, preventing hepato-steatosis (24). This influence of gp130 activation was further demonstrated to be a vital signal mediator in exercise induced weight reduction (25, 26).

Indeed, in our mechanistical investigation, we observed an increase in intracellular lipid accumulation when macrophages were treated with gp130 inhibitor, as evidenced by increased cytoplasmic granularity and neutral lipid content, suggesting a direct impact of gp130 inhibition on lipid uptake and storage mechanisms. This is further corroborated by the upregulation of the PLIN2 gene, a marker of lipid droplet formation. To further identify the intricate of this lipid induction, we conducted a broad gene analysis of for different lipid processing genes. Even though all lipid uptake and shuttle genes showed significant upregulation under the stimulation of oxLDL, an interesting notable increase in LDLR expression under gp130 inhibition regardless of oxLDL supplementation hinted at a possible enhanced cholesterol uptake capacity. Moreover, similar observation was found in the expression of FABP5. Several studies have implicated both FABP4 and FABP5 in atherosclerotic lesions formation (27-29). However, it is very interesting to note that while FABP4 was only marginally detected in early and advanced lesions, the presence of FABP5 is found to be expressed abundantly in these lesions. This increase in expression of FABP5 is also found in areas mostly restricted to infiltrated foam cells (30). Furthermore, the association between FABP5 in particular and macrophage cholesterol efflux capacity (31) further supported the impact of gp130 inhibition on cholesterol regulation.

The absence of significant changes in ACACA expression, coupled with the observed reductions in FASN and DGAT gene expression, indicates a dampened *de novo* lipogenesis and triglyceride production under gp130 inhibition. This is consistent with a cellular attempt to limit further fatty acid synthesis in response to heightened lipid influx.

Another interesting observation we had was the influence of gp130 inhibition on downregulating genes involved in β-oxidation, such as ACADL and CPT1A/CPT2. In a recent work presented by *Zheng et al*, it was demonstrated that the influence of Atorvastatin; a statin medication used for the control of cholesterol levels, induced cardiac pleiotropic effects through inactivating p-STAT3, a transcription factor downstream of gp130 causing the inhibition of CPT1 signaling and thus inhibiting fatty acid oxidation (FAO) (32). Even though the authors believe that this effect holds a protective impact in preventing cardiac hypertrophy and dysfunction, it also further supports a state of altered lipid utilization and metabolism that is specifically tailored towards cholesterol homeostasis.

Paying more attention to the influence of gp130 on cholesterol homeostasis in the presence and absence of oxLDL supplementation, we conducted further characterization analysis of key genes involved in cholesterol *de novo* biosynthesis, storage, and exportation. The activation of the membrane-bound transcription factors SREBPs in both cholesterol synthesis as well as fatty acid synthesis has been well characterized, beside them being a driven factor for both cholesterol and fatty acid production, they also provide the mean by which cellular cholesterol exerts negative feedback on cholesterol synthesis (33, 34). It is not surprising to observe a reduction in SREBF1 and SREBF2 gene expression under the stimulation of oxLDL when compared to vehicle control. However, the use of SC144 to inhibit gp130 activity induced a sustained elevation in SREBF2 that was found not to be reduced under oxLDL stimulation. We believe that this sustains upregulation in SREBF2 subsequently aided the upregulation seen MVK gene expression consequently producing more HMG CoA reductase protein. Nonetheless, as we noticed this upregulation of cholesterol biosynthesis induced by the inhibition of gp130, we also observed strong inhibitory effect seen on ABCA1 at both its gene and protein levels. This observation suggests that under gp130 inhibition this observed intracellular lipid accumulation is more likely to be caused by cholesterol retention. Indeed, cholesterol immunofluorescence staining with Filipin III further defines this impact of cholesterol retention within gp130 inhibited cells. However, this impact was further defined when we investigated the mechanistic ability for THP-1 transformed macrophages to efflux cholesterol under oxLDL stimulation, we found that their ability was significantly abolished when treated with SC144 inhibitor. Translating these findings into a primary human model, we observed that the ability of lean individuals to probably efflux cholesterol from there macrophages indeed seems to be dependent on gp130 activation, as when we pre-treated lean individuals PBMCs with SC144 we observed significant reduction in these cells’ ability to efflux cholesterol to a point where they became similar to obese individuals isolated cells and most likely due to similar impact identified in our THP-1 model.

In conclusion, the findings of this study shed light on the intricate dynamics of IL-6 receptors and their potential role in modulating human macrophage cholesterol homeostasis. By unraveling these molecular intricacies, this research lays the groundwork for future therapeutic interventions aimed at mitigating the progression of atherosclerosis and related cardiovascular diseases.

## Data Availability

The datasets generated and/or analyzed during the current study are not publicly available but are available from the corresponding author on reasonable request.

## Acknowledgements

This work was supported by Kuwait Foundation for the Advancement of Sciences Grant RA CB-2023-017 (FAR.)

## Contributors

FAR. conceived idea, acquired funds, designed study, conducted experiments, data curation, analyzed data, interpreted the results and wrote the manuscript. AAS. conducted experiments, data curation, analyzed data, interpreted the results, and participated in writing the original draft of the manuscript. NAM. participated in conducting experiments, data collection, data analysis, data interpretation and wrote original draft of the manuscript. FAM. participated in study designing, data interpretation, review & editing manuscript. RA. conceived idea, participated in study designing, interpreted the results, acquired funds, reviewed & editing manuscript. All authors contributed to reviewing the paper and all authors have read and approved the final version for submission.

## Data sharing statement

Summarized data are available in the main text or the supplementary materials. There are no restrictions on material or data.

## Disclosure instructions

During the preparation of this work the author(s) used QuillBot AI to improve readability and language. After using this QuillBot AI, the author(s) reviewed and edited the content as needed and take(s) full responsibility for the content of the publication.

